# Correlation between faecal microbial taxa and ulcerative colitis in different phases of disease activity in a north Indian cohort

**DOI:** 10.1101/2021.12.12.21267614

**Authors:** Garima Juyal, Ajit Sood, Vandana Midha, Arshdeep Singh, Dharmatma Singh, Ramit Mahajan, Vijay Verma, Rakesh Bhatnagar, Mohan C Joshi

**Author notes:** Department of Genetics, University of Delhi South Campus, Benito Juarez Marg, New Delhi, India (Garima Juyal). North Florida Research and Education Centre (NFREC)/Institute of Food and Agricultural Sciences (UF/IFAS), University of Florida, Quincy 32351, FL, USA.

## Abstract

**Background and Aims:** A link between gut microbiota and Ulcerative colitis (UC) has been established in several studies. However, only a few studies have examined specific changes in microbiota associated with different phases of disease activity in UC. In this study, we attempted to identify differentially abundant taxa between (i) UC patients and healthy controls and (ii) different phases of disease activity, in a genetically distinct north Indian (NI) population.

**Methods:** 16S rRNA (V3–V4) sequencing of 105 patients with UC [newly diagnosed (n=14); patients in remission (n=36) and active disease (relapse, n=55)]; and 36 healthy controls was performed and analysed.

**Results:** Lower gut microbial diversity; enrichment of lactate-producing bacteria namely *Streptococcus, Bifidobacterium* and *Lactobacillus*; and depletion of butyrate-producing bacteria (e.g., Lachnospiraceae and Ruminococcaceae), were observed among UC patients. Subgroup analysis revealed differential abundance of *Escherichia-Shigella, Streptococcus, Enterococcus* and *Faecalibacterium* in newly diagnosed UC patients. No discrete microbial features were observed between patients in remission and those with active disease. Co-occurrence network analysis revealed a mutualistic association between opportunistic pathogens and *Bifidobacterium* and *Lactobacillus* and antagonistic relationship with butyrate-producers.

**Conclusion:** This first faecal microbiome study among NI UC cohort elucidated dysanaerobiosis; loss of short chain fatty acid producers and enrichment of inflammation associated microbes; population specific differential microbial genera; and a potential microbial signature for early dysbiosis.

## Introduction

Historically considered to be a disease of the western world, ulcerative colitis (UC), has emerged as a global disease ^1^. The etiology of UC is multifactorial. However it is believed that a complex dynamic interplay between genetic factors, environmental influences and intestinal microbiota drives chronic inflammation, characteristic of UC ^2,3^. The conventional treatment strategies focus on inducing and maintaining remission and preventing disease related complications by targeting the dysregulated immune system ^4^. In the past decade however, the intestinal microbiota has unfolded as a crucial integrant in the pathogenesis of UC. Novel microbiota targeted therapies therefore have made inroads into the current treatment paradigms as alternative/adjunctive therapies ^5,6^.

Several microbiome studies, primarily from Western countries, have demonstrated differences in the faecal microbial profile between UC patients and healthy individuals ^7^. However there is paucity of data from developing countries. In light of the differences in genetic predisposition to IBD across ethnically divergent populations ^8^, and their interactions with non-genetic factors predominantly diet, on the development and clinical spectrum of UC, it is imperative to survey the gut microbiome from genetically, culturally and socially divergent populations.

In the current study, we analysed the faecal microbial composition in UC patients and healthy controls of north Indian (NI) origin. Since UC is an emerging disease in developing countries, in contrast to the western world where it has reached a plateau (prevalence equilibrium), we propose that characterizing the gut microbiome of a UC cohort from a developing country like India provides a unique opportunity to identify early microbial markers.

## Materials and Methods

### Cohort

Consecutive patients (age >18 years) with an established diagnosis of UC ^9^ attending the outpatient clinic at Dayanand Medical College and Hospital, Ludhiana, India between January 2016 and December 2017 were categorized into two groups i.e. **newly diagnosed** [patients with active disease (defined as Mayo clinic score >2) diagnosed within four weeks of enrolment with no prior exposure to treatment for UC (including corticosteroids, 5-aminosalicylates (5-ASA), immunomodulators and biologics)]; and **previously diagnosed** (diagnosed >6 months prior to enrolment and on standard of care pharmacotherapy for UC). Patients in this group were further divided into two sub-groups: **relapse:** previously diagnosed UC with active disease and **remission:** previously diagnosed UC in clinical remission (defined as Mayo clinic score ≤2, with each sub-score ≤ 1 and endoscopic sub-score of 0). In patients with clinical symptoms of active disease, a limited sigmoidoscopy was performed for endoscopic evidence of disease activity. Infections like *Clostridioides difficile*, cytomegalovirus (CMV) and/or Epstein Barr virus (EBV) were tested in patients with active disease. Clinical details including patient demographics, disease characteristics and treatment history were recorded. Patients with age <18 years, Crohn’s colitis, indeterminate colitis, *C. difficile* infection, co-infection with CMV or EBV, co-morbid illnesses such as severe heart, lung, or neurological disease, and active malignancies, antibiotic use during the month before inclusion and diagnosis of UC between 4 weeks to 6 months prior to enrolment were excluded. Adult (age >18 years), unrelated volunteers who had no comorbidities or disorders known to be associated with changes in gut microbiota, served as **healthy controls**.

### Faecal DNA extraction and 16S rRNA sequencing

Faecal samples were collected from patients and healthy controls at the hospital settings. The samples were labelled using unique patient specific codes, homogenized and stored at -80°C within 2 hours. DNA was extracted from faecal samples as a non-invasive proxy for the gut microbiome using DNeasy PowerLyzer PowerSoil Kit **(Qiagen/MO BIO cat# 12855-50)** as per manufacturer’s protocol. For profiling microbiome composition, the V3-V4 hyper-variable region of the bacterial 16S rRNA gene was sequenced on Illumina Miseq platform using a commercial facility.

### Data analysis

Paired-end reads were merged using FLASH ^10^ with a minimum and maximum overlap of 30bp and 250bp respectively. Low base quality (average Q≥34) reads were removed using fastx toolkit 0.0.13 **(http://hannonlab.cshl.edu/fastx_toolkit/)**. De-replication was performed in *VSEARCH* ^11^ for the identification of unique sequences. A *de novo* approach in *VSEARCH* ^11^ was used to remove chimeric sequences. Bacterial community composition of a sample was achieved using closed_reference Operational Taxonomic Unit (OTU) picking method in UCLUST ^12^. In this method sequencing reads were clustered based on 97% similarity to form OTU and subsequently one representative sequence from each OTU was picked. Any OTU that had a count of 1 in a single sample (identified as singletons) was removed. Taxonomy assignment was done using RDP classifier ^13^ against Silva-132 database at 97% similarity. OTU table was then normalized to the same depth. Alpha and Beta diversity indices were estimated and distance metrics were represented as principal coordinate analysis (PCoA) plots using QIIME1.9.1^14^. Hierarchical clustering and Co-occurrence network analysis were performed using ClustVis ^15^ and SParse InversE Covariance Estimation for Ecological Association Inference (SPIEC-EASI) program in R ^16^ respectively. Both these methods are detailed in **Supplementary text**.

Shapiro–Wilk normality test was used to test the distribution of data. The bacterial numbers were transformed into relative percentages for statistical analysis. Significant differences in alpha and beta diversity were tested using Mann-Whitney T-test and PERMONOVA respectively. Significant differences in the relative abundance between groups were computed using Mann-Whitney test and p<0.05 was considered statistically significant after Bonferroni correction.

## Results

### Patient characteristics and 16S rRNA amplicon sequencing data

A total of 105 UC patients comprising three subgroups i.e. newly diagnosed, n=14; Relapse, n=55; Remission, n=36 and 36 healthy controls were included in the study **(Table 1)**. Sequencing of V3-V4 region of faecal DNA samples yielded a total of 114200323 ± 478237.89 (sum ± SD) paired-end reads with a read length of 268.37 ± 23.80 (mean ± SD) **[Table 2a]**. After removing low quality reads, chimera sequences and singleton, a total of 300467 ± 221183.15 (mean ± SD) and 458981.08 ± 112054.93 (mean ± SD) sequencing reads were obtained in UC patients and healthy controls, respectively **[Table 2a]**. The sequence reads were normally distributed across samples (**p<0.001**) **[Table 2b]**.

### Microbial composition in UC patients differs from that of controls

Sequencing reads were rarefied to 20,000 sequences per sample to control for variations in sequencing depth. Significant low alpha diversity was observed in UC patients compared to healthy controls [Shannon Index (**P**_**corrected**_**<9.7E-14);** Chao1 (**P**_**corrected**_**<3.5E-14);** number of observed species (**P**_**corrected**_**<3.5E-14) [Figures. 1a-c]**. Beta diversity showed significant dissimilarity in the compositional structure of bacterial communities between UC patients and healthy controls (**PERMONOVA p=0.001; Figure 2)**. Tight clustering in healthy controls indicated high gut microbial compositional similarity among themselves compared to UC patients **[Figure 2a]**. However, a scattered distribution among UC represented heterogeneity underlying disease pathogenesis **[Figure 2b]**. No differences in diversity was observed among the three UC subgroups. However, significant alpha diversity differences were observed between the three UC subgroups and healthy controls **[Supplementary Figs. 1-2]**.

**Figure. 1.**
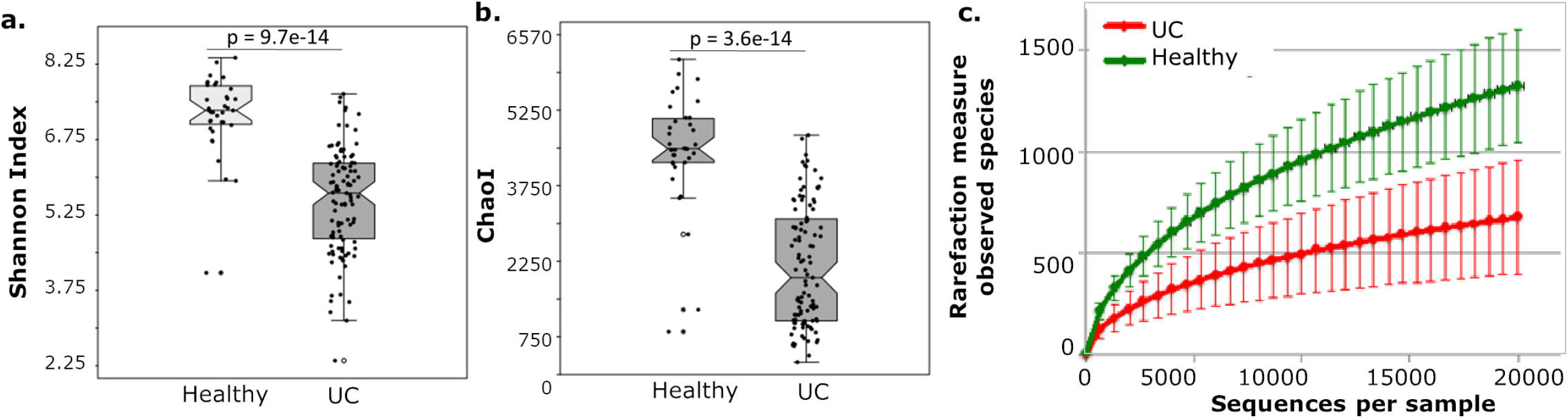
Alpha diversity metrics of fecal bacterial communities in UC and healthy controls. Whisker box plots illustrate (A) species diversity; and (B) species richness between the UC patients and healthy controls. Rarefaction curve (C) shows the observed species at various sequencing depths in UC patients and healthy controls. * represents Bonferroni corrected Mann-Whitney p value.

**Figure. 2.**
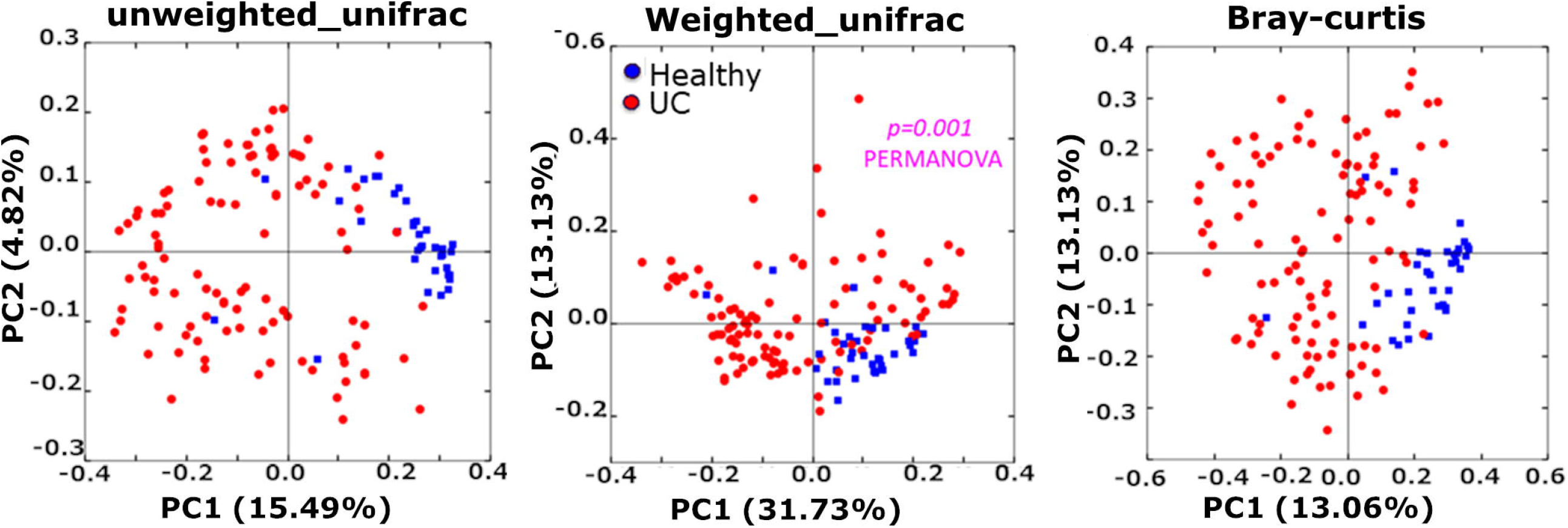
Beta diversity analysis based on the overall structure of the faecal microbiota of UC patients and healthy controls. Principal Coordinate Analysis (PCoA) of UC patients and healthy controls based on (A) unweighted and (B) weighted UniFrac metric and (C) Bray Curtis dissimilarity. Percentages on the x-axis and y-axis correspond to the percent variation in gut microbiome compositions explained by PC1 and PC2. The gut microbiota of UC patients show significant dissimilarity from healthy controls (p=0.001 PERMONOVA).

Overall distribution of predominant phyla that contributed to >99% of the total taxonomy based on median abundance in UC patients and healthy controls is depicted in **Figure 3a**. The most abundant phylum in healthy controls was Firmicutes which accounted for 58.4 ± 8.1% (median abundance ± SD) followed by Bacteroidetes (21.8 ± 9.6%), Actinobacteria (9.1 ± 10.9%) and Proteobacteria (2.1 ± 4.1%). On the other hand in UC cohort, Firmicutes contributed 57.7 ± 19.9% followed by Actinobacteria (18.3 ± 14.0%), Bacteroidetes (5.14 ± 20.4%) and Proteobacteria (1.7 ± 11.4%) **[Figure 3a]**. Of these, the relative abundance of Actinobacteria, Bacteroidetes and Tenericutes significantly differed **(P**_**corrected**_**<0.05)** between UC patients and healthy controls **[Figure 3b]**. When compared independently with healthy controls, Bacteroidetes and Actinobacteria were found significantly altered across the three UC subgroups. In addition, significant difference in the abundance of Bacteroidetes **(P**_**corrected**_**=0.05**) was observed between newly diagnosed and remission groups **(Figs. 4a-b)**. At the genus level, we observed 11 genera in UC patients and 20 genera in healthy controls representing >1.0 % of the total OTUs **(Figure 5)** which reiterates higher diversity and richness in the heathy group. Among these, relative abundance of 18 were significantly altered **[P**_**corrected**_**<0.05]** in UC patients compared to healthy controls **[Figure 6a]**. In the subgroup analysis, we observed nine genera in newly diagnosed, 13 in remission and 11 in relapse which represented >1.0 % of the total OTUs **[Figure 5b]**. Of these, only four namely uncultured_Bacteroidetes, *Escherichia-Shigella, Enterococcus*, and *Ruminococcaceae* were significantly altered **[P**_**corrected**_**<0.05]** across the three subgroups **[Figure 6b]**. Furthermore, on comparing the three UC subgroups independently with healthy controls, 19 genera were found to be significantly altered (**P**_**corrected**_**<0.05**) **(Figure 6b)**. The p-value of altered taxa and their abundance are presented in **Table 3**.

**Figure. 3.**
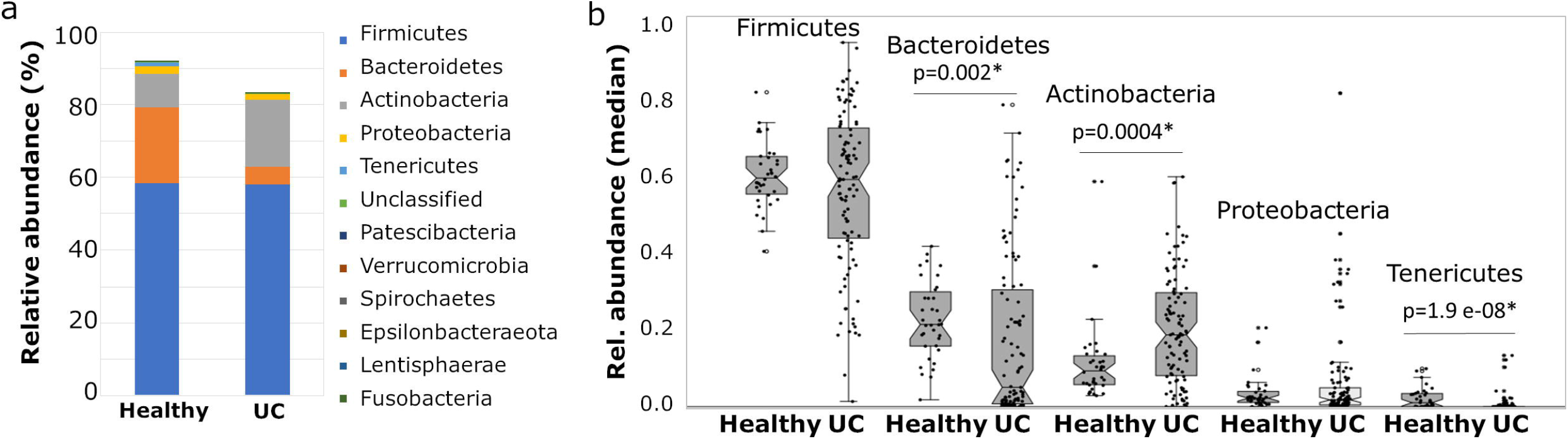
Taxonomic diversity of faecal microbiota composition at the phylum level among UC patients compared to healthy. (A) Distribution of faecal microbiota composition and (B) Inter-individual variation of predominant phyla based on their relative abundance. * represents Bonferroni corrected Mann-Whitney p value.

**Figure. 4.**
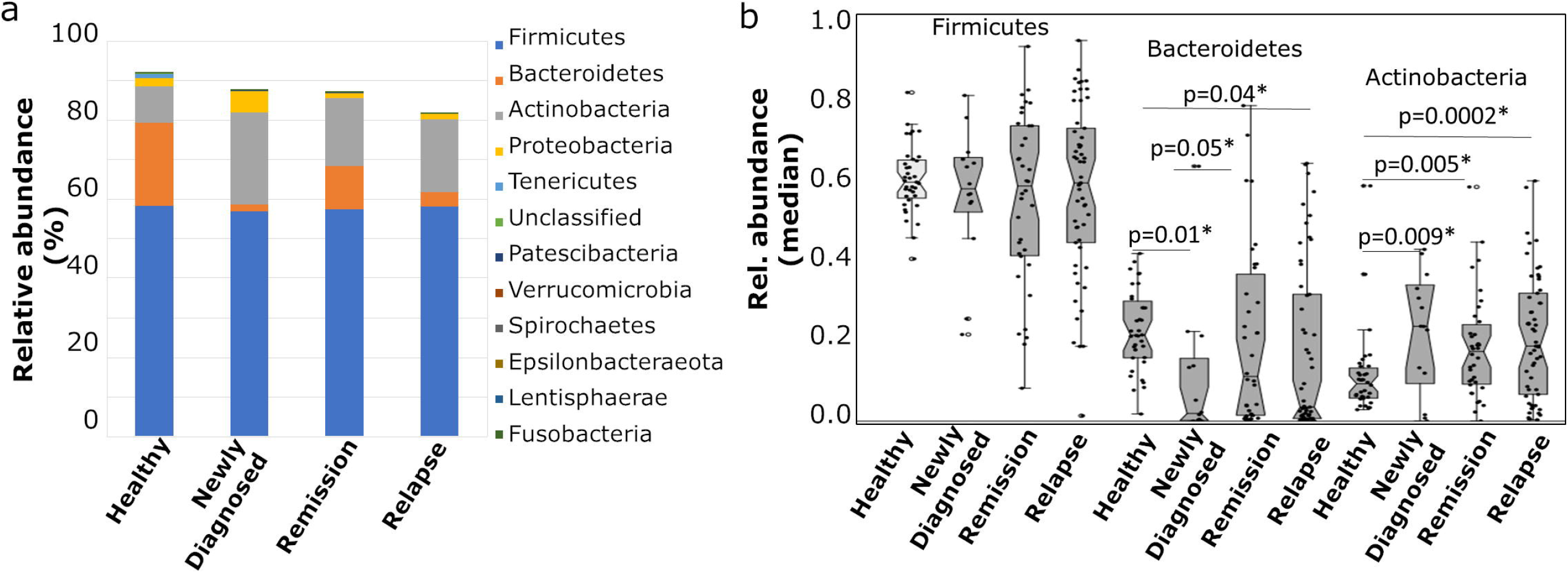
Taxonomic diversity of faecal microbiota composition at the phylum level within UC subgroups compared to healthy. (A) Distribution of faecal microbiota composition and (B) Inter-individual variation of predominant phyla based on their relative abundance. * represents Bonferroni corrected Mann-Whitney p value.

**Figure. 5.**
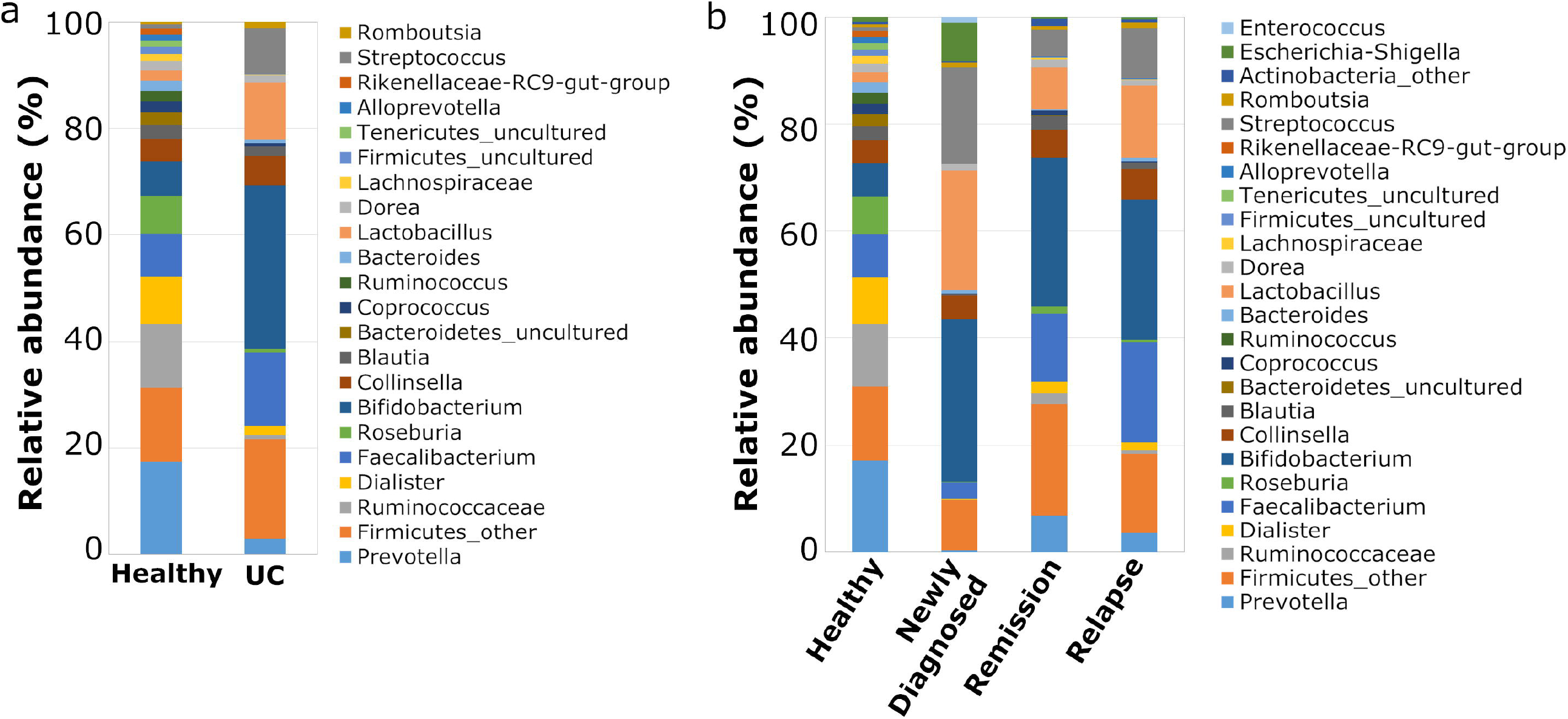
Taxonomic diversity of faecal microbiota composition at the genus level. (A) total UC patients and (B) UC subgroups compared to healthy * represents Bonferroni corrected Mann-Whitney p value.

**Figure. 6.**
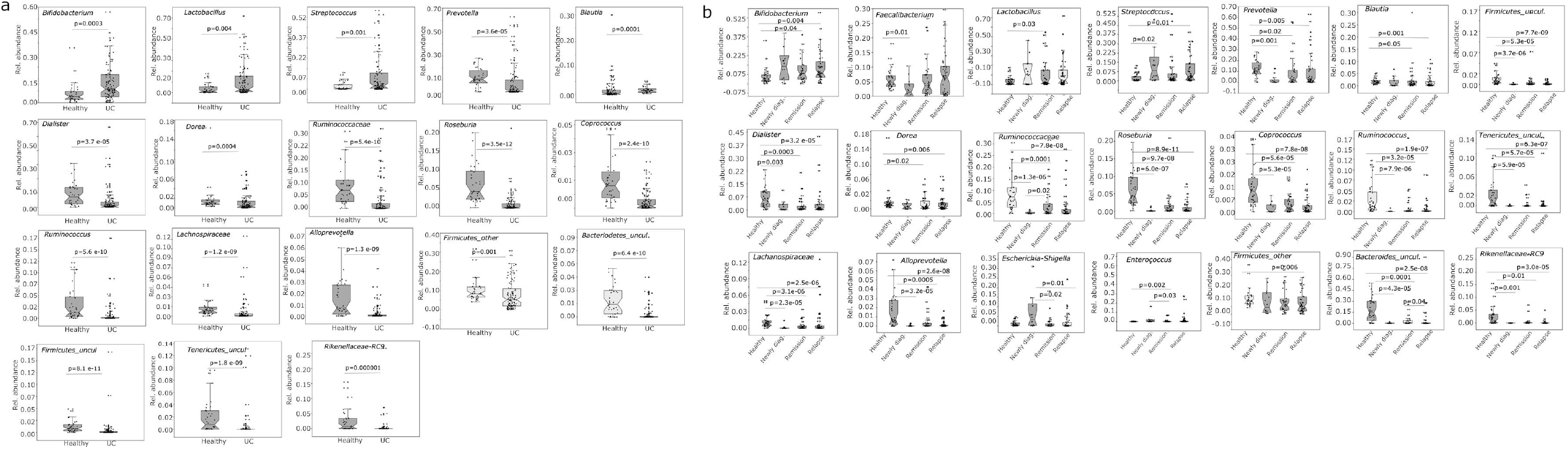
Differential abundances of microbial genera representing >1.0% of total OTUs. (A) UC patients and (B) UC subgroups compared to healthy. * represents Bonferroni corrected Mann-Whitney p value.

To validate the differential abundance of nitrate reducing bacteria in the faeces of UC patients and healthy controls nitrate reduction activity was determined using nitrate broth **[detailed in supplementary text]**. Significant increase in the number of nitrate reducing bacteria per gram of faeces was observed in UC patients compared to healthy controls **(P**_**corrected**_**=E-08)**. However, no differences were observed across the three UC subgroups **[Supplementary Figure 3]**.

### Hierarchical clustering reveals newly diagnosed UC as a unique clad

Clustering analysis categorized UC patients and healthy controls into distinct clads based upon differential OTU abundance (>1%) at genus level and heat maps are represented in **Fig 7a**. Barring four, the remaining genera were found to be statistically significant in the abundance analysis **[Table 3]**. Further, cluster analysis also illustrated that newly diagnosed UC patients formed a distinct clad compared to remission, relapse and healthy controls **[Fig 7b]**.

**Figure. 7.**
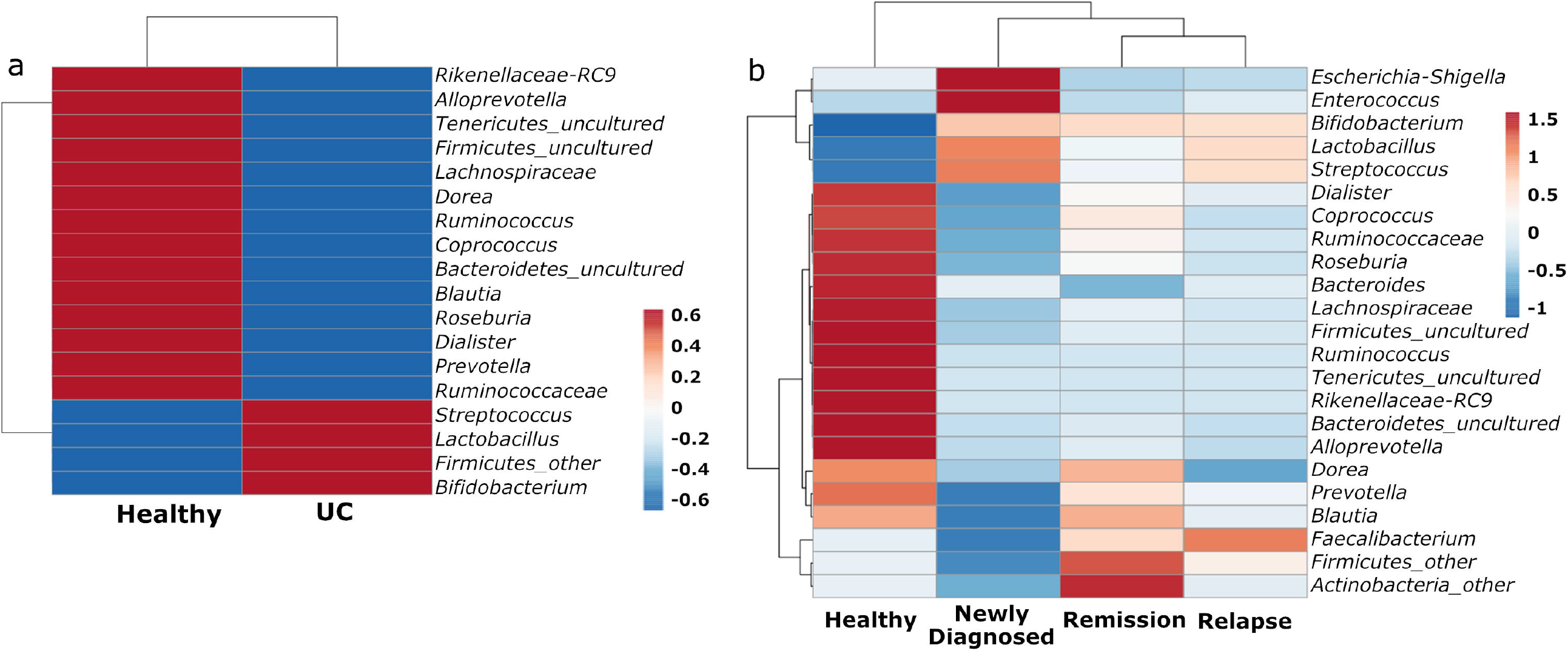
Hierarchical clustering analysis of genera representing >1.0% of total OTUs. (A) all UC patients and (B) UC subgroups compared to healthy. The color code indicates relative abundance of OTUs ranging from red (high) to blue (low). The OTUs were transformed to (ln(x + 1)) values, rows were centered and vector scaling was applied using ClustVis.

### Co-occurrence network analysis reveals enrichment of pathobionts in UC patients

A total of 115-116 pruned OTUs were included for co-occurrence network analysis to study their potential relationships in different pathological contexts among UC patients. Such analysis gives an initial cue on the structure of the community and how metabolite cross-feeding/metabolic dependency and nutritional preferences contribute to microbial assemblage and their maintenance. This analysis revealed (a) *Streptococcus, Escherichia-Shigella, Bifidobacterium, Lactobacillus* and *Faecalibacterium* in newly diagnosed; (b) *Streptococcus, Lactobacillus, Prevotella* in relapse; (c) *Streptococcus, Blautia* and *Erysipelotrichaceae* in remission; and d) *Ruminococaccae, Bifidobacterium, Prevotella* and *Bacteroides* in healthy controls, to be the predominant taxa in the networks **[Figure 8]**.

**Figure. 8.**
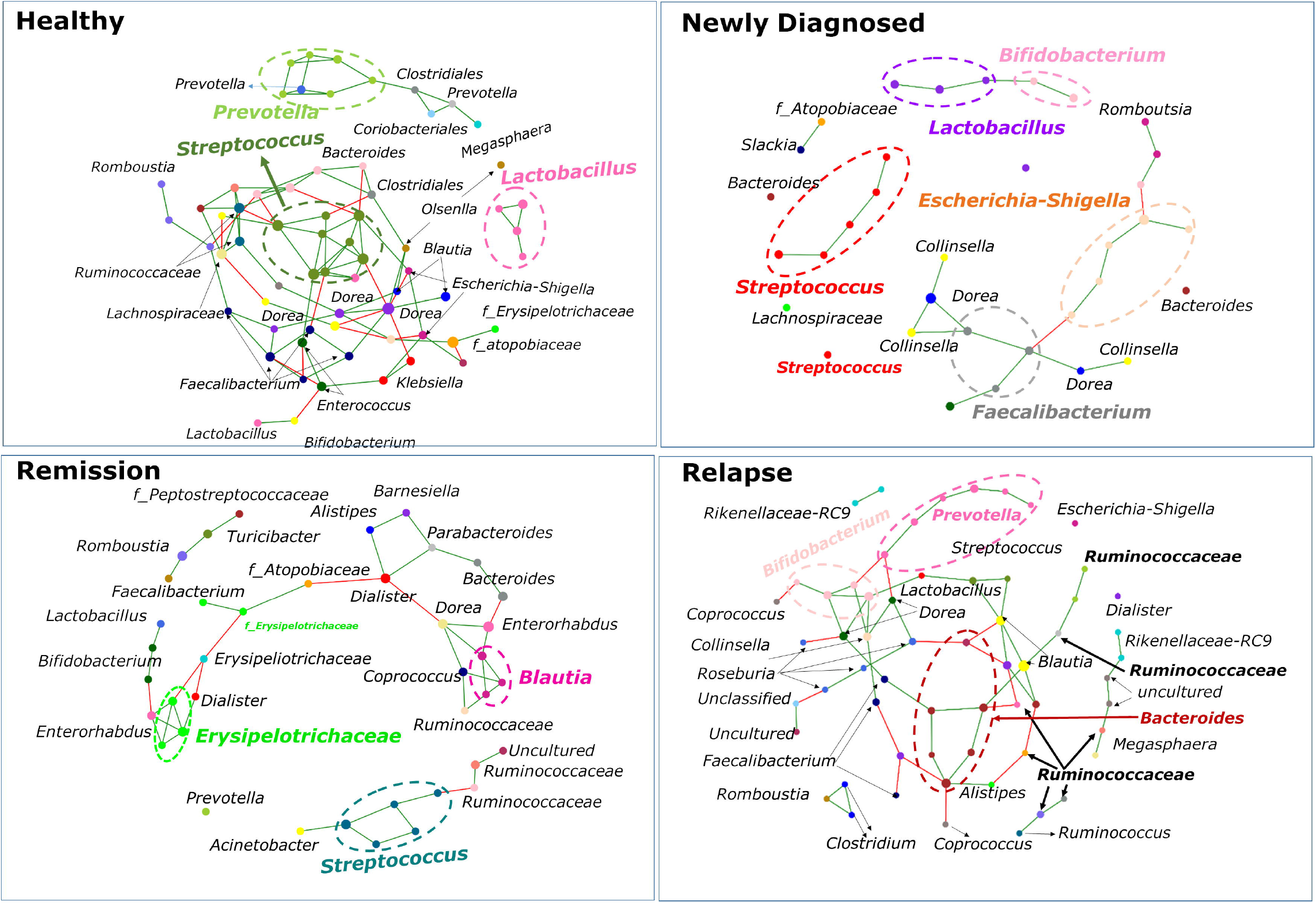
Co-occurrence networks analysis of microbial genera based upon its relative abundance. (A) Newly Diagnosed, (B) Relapse, (C) Remission and (D) Healthy. Each node represents genus and is uniquely colored. The size of each node is proportional to the abundance of the genus in log scale. Line between two nodes represents correlation; green and red lines indicate positive and negative association respectively. Predominant network in UC subtypes and healthy controls is represented with dotted circle.

## Discussion

Gut microbial dysbiosis in terms of alteration in composition, reduction in biodiversity and consequently loss of metabolic and immune homeostasis has been associated with development of UC ^17^. To date, most of the published studies on UC from India have been largely based on either quantification of specific bacteria in tissue biopsy or faecal samples ^18–20^ or 16S rRNA analysis of limited tissue biopsy samples ^19,21^. To our knowledge, this report is the first description of faecal microbiota profile of NI UC patients which provides a snapshot of the alterations of the microbiota in different phases of disease.

We report that the ethnically distinct NI UC population exhibited a marked reduction in the bacterial diversity and significant alteration in the structure of faecal bacterial community as compared to healthy controls **[Figures. 1 & 2]** lending support to the well noted central feature of dysbiosis in UC pathology. We found significant loss of genera comprised of dominant anaerobes alongside expansion of subdominant facultative anaerobes suggestive of a disruption in anaerobiosis in the gut lending support to oxygen hypothesis ^22^, discussed in the later section. Each of these genera has been discussed below in detail. Findings from subgroup analyses have been discussed in the subsequent section.

### UC patients vs healthy controls

#### Loss of SCFA producing taxa

We observed a significant loss of anaerobic SCFA producing bacteria in UC patients compared to healthy controls **[Table 3; Figure 6a]**. This is in consonance with previous studies from European and Asian populations ^7,23,24^. SCFAs are organic products mainly composed of acetate, propionate, and butyrate produced after fermentation of dietary fibers and resistant starches in the colon. They act as signaling molecules and regulate different biological processes, including the promotion of gut integrity, immune response, reduction of pathogenic bacterial population; and prevention of unwanted infiltration of bacteria from lumen to lamina propria ^25,26^. This suggests that lower abundance of SCFA-producing bacteria in UC patients may favour a shift towards an inflammation-promoting microbiome, thereby enhancing host inflammation and modifying the disease behaviour. This is in line with oxygen hypothesis which posits that chronic inflammation of intestinal walls results in extravasation of blood from ulcerated tissue which leads to increased release of hemoglobin carrying oxygen and reactive oxygen species into the intestinal lumen, which in turn creates a microenvironment that favors expansion of facultative anaerobes. The resulting decrease in anaerobes (which release anti-inflammatory compounds) leads to increased inflammation, establishing a positive feedback loop that accelerates the disease process ^22^.

#### Enrichment of potentially harmful bacteria

A significant enrichment of *Streptococcus*, a lactic acid-producing bacteria (LAB) was observed in UC patients **[Table 3, Figure 6]** which is in line with earlier reports ^27–29^. Some species of this bacterium have shown to induce or promote pro-inflammatory response in macrophages, human tissue, and intestine ^29^ suggesting that metabolites and structural components of *Streptococcus* and immune cells are at direct/indirect interplay and lead to excessive inflammation. In a recent study, increased abundance of f_Streptococcaceae among IBD patients was correlated with proton pump inhibitors (PPIs) ^3,30^. However, in our case none of the UC patients were using PPIs suggesting other factors responsible for *Streptococcus* abundance. Interestingly, evidences are now emerging and pointing to association between higher levels of gut inflammation in IBD patients and increase in bacteria typical of the oral cavity ^28^. Although oral microbes are innately resistant to colonization in healthy guts, it has been proposed that inflammation or strain-specific adaptation (including antibiotic resistance and virulence genes) of oral microbes allows their colonization in the gut resulting in exaggerated inflammatory response ^31^. Therefore, it would be interesting to further explore the link between oral microbiome and UC.

The other enteric anaerobes which were found to be enriched in UC patients were also LABs namely *Bifidobacterium* and *Lactobacillus* **[Table 3, Figure 6]**. These findings are in stark contrast to majority of earlier studies from west ^7,23,32^. However, enrichment of these bacteria have been reported in the faecal and tissue biopsy samples of active UC patients from Chinese and Russian populations ^33,34^. Such observed differences reinforce the potent role of host genetics in determining the composition of the gut microbiota. This derives support from recent reports documenting a) that differences in microbiota are highly impacted by the ethnic background ^35^; b) microbiota is partly heritable as evidenced in studies analysing both concordant and discordant twins for IBD ^36^; c) influence of genomic loci on several microbial genera in mice ^37^; and d) specific variants of the *NOD2* associated with changes in the abundance of the f_Enterobacteriaceae in IBD patients ^38^. Importantly, evidences now emerging pinpoint that these commensals have the potential to become opportunistic or promote colonization of other pathobionts and elicit immunostimulatory effects ^39,40^. For instance, a recent study in mice demonstrated that the introduction of a single commensal species, *Lactobacillus reuteri*, was sufficient to exacerbate experimmental autoimmune encephalomyelitis in a genetically susceptible host ^41^. This suggests that these “good” commensals can become “bad” and can cause unwarranted immune stimulation in genetically susceptible individuals reiterating the likely effect of host genetics on microbiome composition. Other than host genetics, differential selection pressure through diet ^42,43^; disease characteristics; and persistent use of 5-ASA (which changes the colonic luminal pH and hence promote growth of *Bifidobacterium* and *Lactobacillus)* ^35^, could be the non-genetic factors that may have contributed on the gut microbial assemblage. Considering the global uniformity of treatment modalities it seems that effect of disease modifying drugs on expansion of these genera is minimal. A few recent studies which have shown that disease activity, 5-ASA dosage or addition of glucocorticoids had no impact on the microbial stability ^44^ lend support to this.

### Subgroup analysis

#### Early dysbiosis hallmark

To avoid the confounding effects of long-standing disease, disease modifying agents (such as medication), and co-morbid conditions which could have potential effect on microbiome composition, treatment naïve newly diagnosed UC patients were analysed in relation to treatment experienced patients [remission and relapse] and healthy controls in this study. An over-representation of *Escherichia-Shigella, Enterococcus* and *Streptococcus* in newly diagnosed UC patients which is in line with prior studies from both adult ^7^ and paediatric ^45^ UC patients was notable **[Table 3]**. *Escherichia-Shigella* and *Enterococcus* have been reported to have the ability to adhere to and invade intestinal epithelial cells, leading to an inflammatory immune response ^46^. Besides other SCFA producing taxa, *Faecalibacterium*, a dominant species in the gut microbiota of healthy subjects and which has anti-inflammatory properties was found to be significantly depleted in newly diagnosed UC patients **[Table 3]**. The reduction of extremely oxygen-sensitive *Faecalibacterium* and increase in facultative anaerobes reiterates increased oxidative stress in lumen (oxygen hypothesis, discussed above). *F. prausnitzii*, the sole species of *Faecalibacterium* has been suggested to be an important regulator of intestinal homeostasis ^47^ and its association with IBD has been established in several gut microbial studies ^7,23^.

Similar to studies investigating paediatric patients with IBD, the association of *Enterococcus, Escherichia, Streptococcus*, and *Faecalibacterium* with newly diagnosed adult UC patients observed in this study may represent an early dysbiotic shift triggering disease. This may imply that these genera could be considered as early non-invasive faecal biomarkers for epithelial dysfunction which includes inflammation, aggravated intestinal injury and increased intestinal permeability.

#### No microbial shift between UC patients in remission and relapse

Gut microbial profiling during different phases of disease activity in UC has been explored in only a few studies with inconsistent and conflicting results ^723^. In the present study, no significant differences were found between disease activity and faecal microbiota (except uncultured_Bacteroidetes) **[Table 3; Figures 6b, 7b]**. Consistent with our observation, a few earlier studies have shown no microbial differences between inactive and active UC patients ^44,48^. Additionally, no significant alteration in the bacterial abundances in relation to disease activity were observed in UC patients of European origin ^3^. Similar trends were also observed in two other independent longitudinal studies wherein gut microbiota in UC patients remained highly stable regardless of disease stage, disease activity or treatment escalation ^44,49^. Thus, associations identified in previous cross-sectional studies may more likely reflect inter-individual variation rather than disease activity. No discrete microbial features between remission and relapse groups may highlight the high level of host genetic and gut microbial heterogeneity. Therefore, to unravel robust shifts in the microbiota related to disease activity species/strain mapping and metabolite profiling may be essential.

#### *Streptococcus*-a persistent pathobiont across the three UC subgroups

No differential abundance of *Streptococcus* was observed across the three UC subgroups. However, on comparing these subgroups independently with healthy controls we observed significant enrichment of *Streptococcus* in newly diagnosed patients and those in relapse **[Table 3]**. Although not statistically significant, *Streptococcus* was found to be higher in remission group compared to healthy **[Figure 6]**. The enrichment of *Streptococcus* across the three UC subgroups **[Figs. 6-8]** suggest that colonization of *Streptococcus* is not transient but resilient (despite treatment regimen or dietary modifications) and thus has a continued role in disease pathology. Of note, the positive correlation of *Streptococcus* with *Lactobacillus, Bifidobacterium* and *Escherichia*-*Shigella* as evident in our co-occurrence analysis **[Figure 8]** suggests mutualistic association among them and validates the relative abundance findings **[Figure 6; Table 3]**.

### Enrichment of nitrate reducing bacteria as an indicator of inflammation

Culture-based analysis showed an increased abundance of nitrate reducing bacteria in the faeces of UC patients compared to healthy controls. These bacteria are capable of converting nitrate to nitrite which are further reduced to nitric oxide and ammonia (nitrogen reactive species). It is now widely believed that oxidants, including free radicals, such as nitric oxide, play a key role in the initiation and perpetuation of inflammation and in the subsequent tissue damage in IBD ^50,51^. No significant differences in nitrate reducing activity was observed among the three UC subgroups, however the activity in each subgroup was found to be significantly higher compared to healthy controls **[Table 3; Supplementary Figure 3]**. While *Escherichia-Shigella* and *Enterococcus* attributed to a higher nitrate reducing activity in newly diagnosed UC patients, *Bifidobacterium* and *Lactobacillus*, which have been recently reported to have nitrate reducing activity ^52^, may explain higher activity among patients in remission and relapse. Interestingly, the latter derives support from a recent study from India wherein significant enrichment of *Lactobacilli* was observed in the faecal samples of active UC patients compared to healthy controls and their level significantly reached close to controls during remission ^19,32^. These observations confirm increased oxidative stress in the gut lumen; and independently validates our observations.

Acknowledging the modest sample size, the significant associations of bacterial taxa (as detailed above) with UC, identified in the present study require replication in larger, independent cohorts of matched ethnicity. However, the high concordance between our findings and several other reports discussed above clearly indicate that our study is well powered. Furthermore, the limitation that 16S rRNA sequence analysis does not enable species/strain assignment, high-resolution mapping of gut microbiome in NI UC patients is highly encouraged. This will unravel more robust disease endotype specific associations driven by select strains which may serve as biomarker. The search for relevant uncultured/unnamed species will expand our knowledge on their relevance in the pathogenesis of UC.

In summary, this first sequencing based faecal microbiome analysis in a NI UC cohort revealed reduced bacterial diversity and altered composition. Most of the genera identified in our study have shown recurring association with UC across ethnically and geographically distinct populations despite differences in methodology, bio-specimens and reference databases used. Our observations may imply that a) restoration of health promoting SCFA producing bacteria could be a pragmatic solution for mitigating disease; and b) *Escherichia-Shigella, Enterococcus* and *Streptococcus* could serve as non-invasive microbial signatures for inflammation of high clinical utility. Higher abundance of *Bifidobacterium* and *Lactobacillus* among NI UC patients suggest inherent population specific differences in disease pathogenesis driven by host genetics, diet and environmental attributes. Their relationship with UC pathogenesis however warrants further investigations. A deeper understanding of how bacterial genera identified in this study are involved in interactions with host genetics, diet and lifestyle factors etc. either by regulating microbial pathways or by production of specific metabolites may facilitate identification of robust prognostic and diagnostic markers and for better microbial based therapeutic interventions.

## Data Availability

All data produced in the present study are available upon reasonable request to the authors

## Ethical consideration

The Institutional ethics review board of Jawaharlal Nehru University, New Delhi and Dayanand Medical College and Hospital, Ludhiana approved the study (IEC numbers 2017/SERB Young Scientist/125 and DMCH/R&D/2015/238 respectively) approved the study. Written informed consent was obtained from each participant.

## Funding

The financial assistance for this work was provided by Science and Engineering Research Board, New Delhi, vide F. NO. SB/YS/LS-191/2014 to GJ.

## Acknowledgements

The authors thank Prof. Thelma BK, Department of Genetics, University of Delhi South Campus for critical discussions throughout the study; MedGenome Labs Ltd. for 16S rRNA sequencing service; Mr. Vikas for logistic support at Dayanand Medical College and Hospital, Ludhiana, India; MCJ acknowledges funding support from DBT/Wellcome Trust Intermediate grant (IA/I/15/2/502086).

## Data availability

The data underlying this article will be shared on reasonable request to the corresponding author.

## Competing interests

The authors declare that they have no competing interests.

## Authors’ contributions

GJ, AjS and VM conceived and designed the study; AjS and VM recruited clinical samples and clinical data; GJ performed faecal DNA isolation, data analysis, interpretation of data, wrote the first draft and secured funding; AS and RM contributed to patient follow-up, clinical data acquisition; DS to sample collection and clinical data acquisition; VV set up the data analysis pipeline and performed independent data analysis; RKB provided infrastructure and other support for conducting the study to GJ as a SERB fellow; MCJ provided critical scientific and technical inputs throughout the study; All authors contributed to manuscript writing and approved the final submitted version.

## Legends

**Table 1. Baseline clinical characteristics of UC patients enrolled in the study**

**Table 2 Sequencing reads obtained a) before and after quality control and b) distribution of sequencing reads after quality control**

**Table 3. Relative abundance of top genera (>1% prevalence) in healthy controls, total UC patients and UC subgroups**

**Supplementary Figure. 1 Alpha diversity metrics of fecal bacterial communities in UC subgroups and healthy controls**. Whisker box plots represent comparison of

(A) species diversity and (B) species richness between the three UC subgroups and healthy controls. Rarefaction curve (C) shows the observed species at various sequencing depths in three UC subgroups and healthy controls. * represents Bonferroni corrected Mann-Whitney p value.

**Supplementary Figure. 2 Beta diversity analysis based on the overall structure of the fecal microbiota between UC subgroups and healthy controls**

Principal Coordinate Analysis (PCoA) of UC subgroups i.e. newly diagnosed (red), remission (blue) and relapse (orange) and healthy controls (green) based on (A) unweighted and (B) weighted UniFrac metric and (C) Bray Curtis dissimilarity. Each point in the plots represents the composition of the faecal microbiome of one individual and distances indicate degree of similarity to other individuals. The closer the spatial distance of the sample, the more similar the species composition of the sample. Percentages on the x-axis and y-axis correspond to the percent variation in gut microbiome compositions explained by PC1 and PC2. No significant differences among the three UC subgroups were observed.

**Supplementary Figure. 3 Relative nitrate reducing activity**

Whisker box plots illustrate relative absorbance (620nm) per gram of faeces observed in nitrate broth of (A) UC patients and healthy and (B) UC subgroups. Each dot represents relative absorbance in a sample. The bottom, middle, and top boundaries of each box represent the first, second (median), and third quartiles of the relative absorbance. The whiskers (lines extending from the top and bottom of the box and ending in horizontal cap) extend to points within 1.5 times the interquartile range. The points extending above the whiskers are outliers. *represents Mann-Whitney Bonferroni corrected p value.

## Notes

### Competing Interest Statement

The authors have declared no competing interest.

### Funding Statement

This study was funded by Science and Engineering Research Board, New Delhi, vide F. NO. SB/YS/LS-191/2014

### Author Declarations

The Institutional ethics review board of Jawaharlal Nehru University, New Delhi and Dayanand Medical College and Hospital, Ludhiana approved the study (IEC numbers 2017/SERB Young Scientist/125 and DMCH/R&D/2015/238 respectively) approved the study.

