## Supplementary figures and images for "Correlation between faecal microbial taxa and ulcerative colitis in different phases of disease activity in a north Indian cohort"

### Supplementary Figure 1

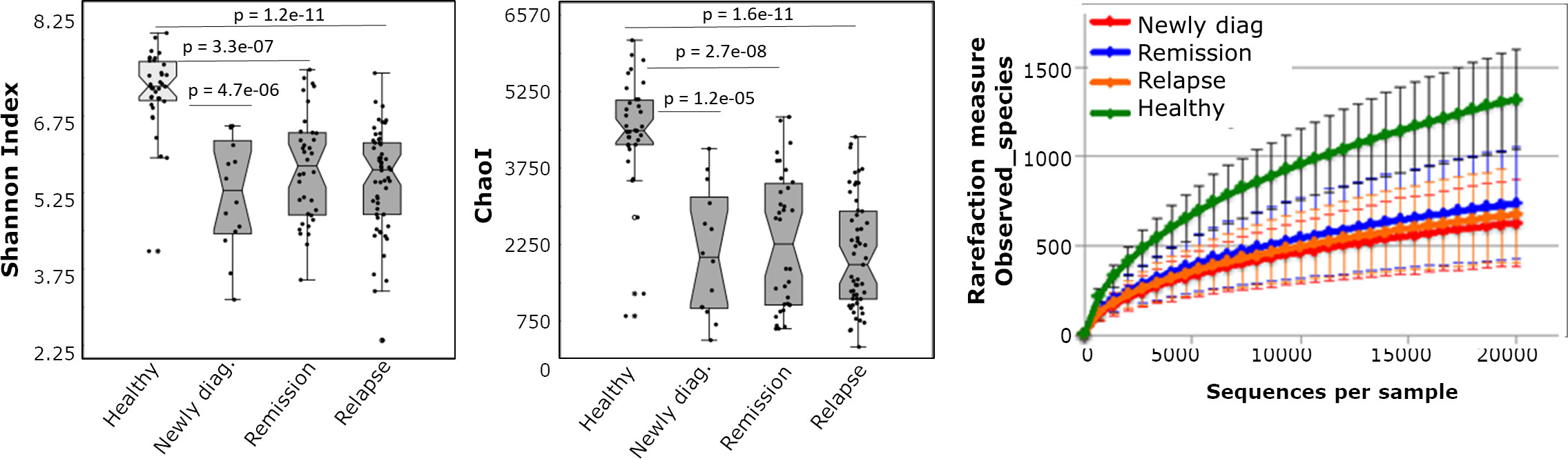

### Supplementary Figure 2

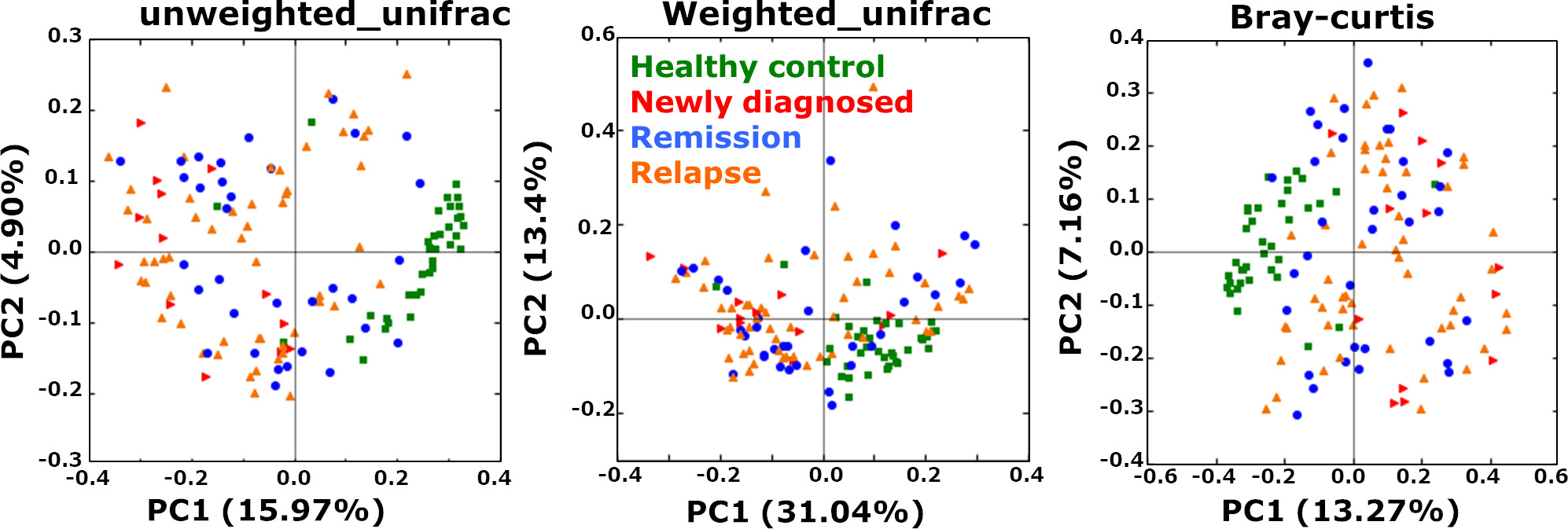

### Supplementary Figure 3

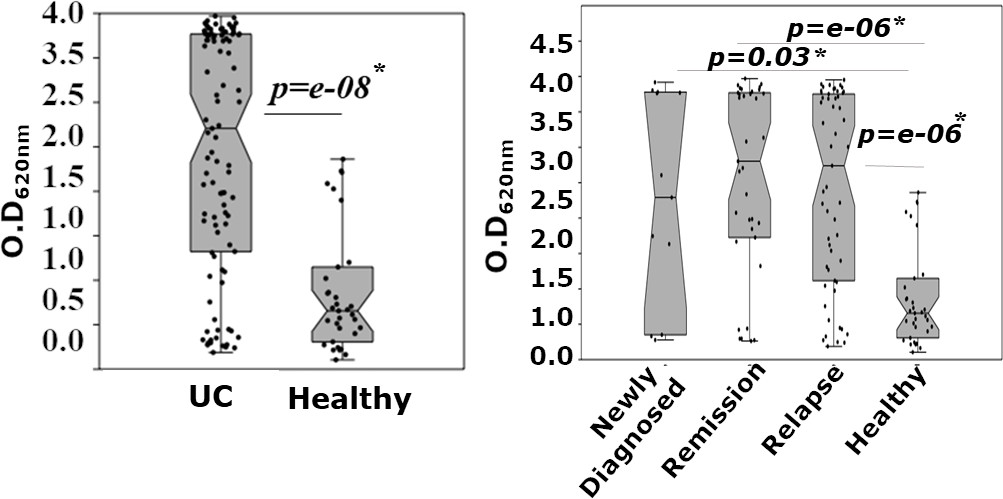
