## Supplementary text for "Correlation between faecal microbial taxa and ulcerative colitis in different phases of disease activity in a north Indian cohort"

**Methods**

**Hierarchical clustering**

Genus representing >1.0% of total OTUs were selected for the analysis. The OTUs were transformed to (ln(x + 1)) to account for zero or negative values, rows were centered and vector scaling was applied. Subsequently, a heatmap was generated based upon correlation distance and average linkage across the three UC sub-groups and healthy controls using ClustVis [27]**.**

**Co-occurrence network analysis**

To explore relationships between the gut microbial taxa in UC patients and healthy individuals, co-occurrence network analysis was performed using SParse InversE Covariance Estimation for Ecological Association Inference (SPIEC-EASI) program in R [28]. It infers an ecological network (inverse covariance matrix) from compositional data using the log-ratio transformation and sparse neighbourhood selection. Briefly, raw biom file was used to prune total OTUs based on their taxa sum (>10,000) and network analysis was performed with 99 permutations on these pruned OTUs. Networks with minimum connections in the range of 2-4 were retained and plotted.

**Nitrate reduction test**

It has been hypothesized that while fighting against pathogenic bacteria host inflammatory response generates nitrate as by-product, which gives an advantage to potentially harmful commensals such as Enterobacteriaceae to bloom as they have an ability to degrade and utilize non-fermentable substrates (nitrate) as energy source for their growth. Since obligate anaerobic microbes such as SCFA producers cannot utilize non-fermentable substrates, it enables for example, *E. coli* to sidestep this competition. Through this mechanism, inflammation contributes to a bloom of nitrate-respiration proficient Enterobacteriaceae, providing a plausible explanation for the dysbiosis associated with intestinal inflammation [29]. To validate this we determined the differential abundance of nitrate reducing bacteria in the faeces of UC patients and healthy controls using nitrate broth assay. Fecal samples of UC subjects and healthy controls were grown in Nitrate broth (HiMedia, cat#M439S). Briefly, faecal samples (0.2 gm) were homogenized in 1x phosphate buffer saline, pH=7.0. Debris were removed by spinning at 12,000 rpm and 100 microliter of supernatant was inoculated into 1ml of nitrate broth. Post 24-48 hours incubation at 37^o^C, 100 microliter of freshly prepared Sulfanilic acid and alpha Napthylhylamine were added to the culture. The change in broth colour (at 620 nm) due to production of nitrite (Griess reaction) reflected/correlated with presence/absence of nitrate reducing bacteria in the sample.
